# The importance of continued non-pharmaceutical interventions during the upcoming SARS-COV-2 vaccination campaign

**DOI:** 10.1101/2020.12.23.20248784

**Authors:** Marta Galanti, Sen Pei, Teresa K. Yamana, Frederick J. Angulo, Apostolos Charos, David L. Swerdlow, Jeffrey Shaman

## Abstract

In this communication we assess the potential benefit of SARS-COV-2 pandemic vaccination in the US and show how continued use of non-pharmaceutical interventions (NPIs) will be crucial during implementation.

## Introduction

A global effort to develop vaccines against SARS-COV-2 began early in 2020, and several vaccine candidates have now reached final stages of the approval process. During mid-December the U.S. FDA is scheduled to discuss emergency use authorization (EUA) for two vaccines that have successfully concluded their phase 3 trials [1]. Vaccine distribution in the US will likely begin shortly after authorization. Both vaccines use mRNA technology and require two doses administered 3 and 4 weeks apart to reach the full 90-95% efficacy [2]. In light of limited supply, the Center for Disease Control and Prevention (CDC) has indicated it may prioritize vaccination for: 1) healthcare workers, 2) long-term care facility residents, 3) other essential workers and 4) people at higher risk for severe illness [3]. In this communication we assess the potential benefit of SARS-COV-2 pandemic vaccination in the US and show how continued use of NPIs will be crucial during implementation.

## Methods

We simulated the progression of the pandemic at the US state level for different combinations of vaccination and other non-pharmaceutical interventions (NPIs), including school/business closure and social distancing. Projections were made with a SEIRV (Susceptible-Exposed-Infected-Recovered-Vaccinated) compartmental model. Simulations were generated for all 50 states with the population stratified by age and priority group. The model was parametrized using the age and population type-adjusted posterior distributions previously estimated with a non-stratified metapopulation model [4] (see **Supplementary Material**). Our vaccination timeline assumed 40 million doses available by the end of December and 10 million additional doses/week thereafter [5] (Figure 1A) distributed to the population according to prioritization guidelines. Vaccination was administered regardless of previous history of infection and was assumed to be 90% effective preventing infection in susceptible individuals 1 week after the second dose. The seven vaccination and NPI scenarios are described in Table 1. We did not account for the possibility of reinfection in any of the simulations.

**Table 1:**
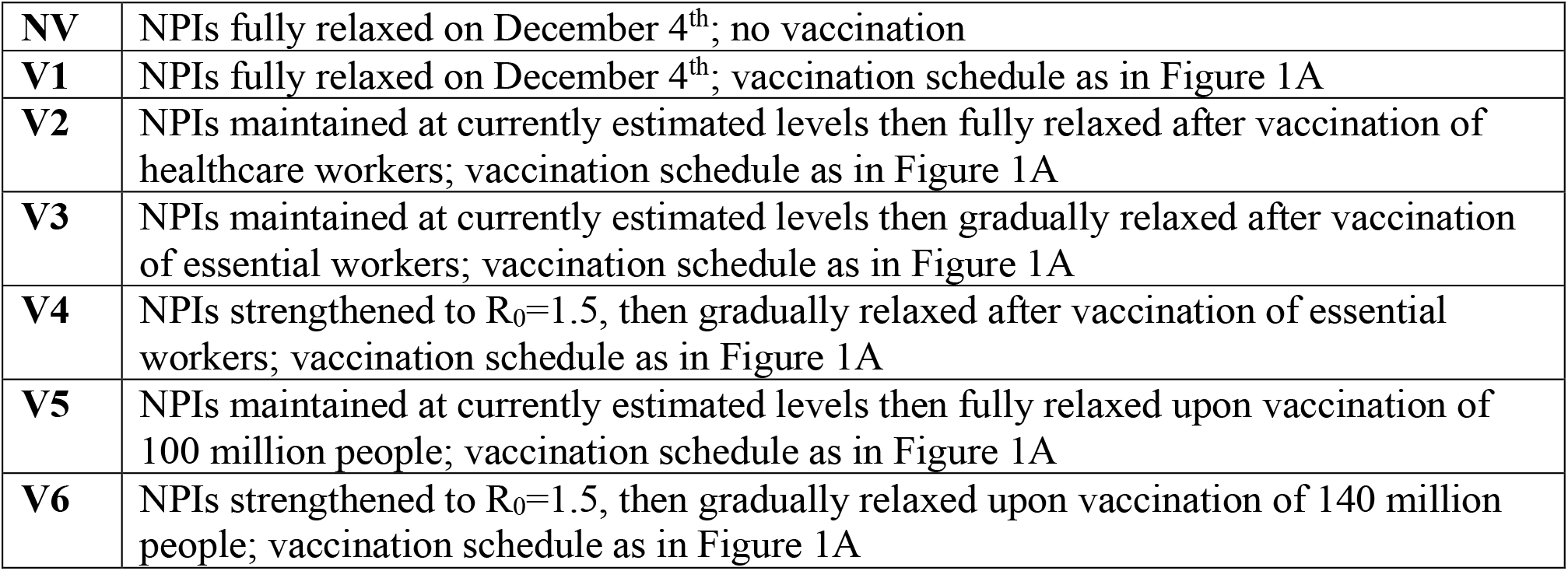
Scenarios for vaccination and NPIs.

|  |  |
| --- | --- |
| **NV** | NPIs fully relaxed on December 4<sup>th</sup>; no vaccination |
| **V1** | NPIs fully relaxed on December 4<sup>th</sup>; vaccination schedule as in Figure 1A |
| **V2** | NPIs maintained at currently estimated levels then fully relaxed after vaccination of healthcare workers; vaccination schedule as in Figure 1A |
| **V3** | NPIs maintained at currently estimated levels then gradually relaxed after vaccination of essential workers; vaccination schedule as in Figure 1A |
| **V4** | NPIs strengthened to $R_0=1.5$, then gradually relaxed after vaccination of essential workers; vaccination schedule as in Figure 1A |
| **V5** | NPIs maintained at currently estimated levels then fully relaxed upon vaccination of 100 million people; vaccination schedule as in Figure 1A |
| **V6** | NPIs strengthened to $R_0=1.5$, then gradually relaxed upon vaccination of 140 million people; vaccination schedule as in Figure 1A |

**Figure 1:**
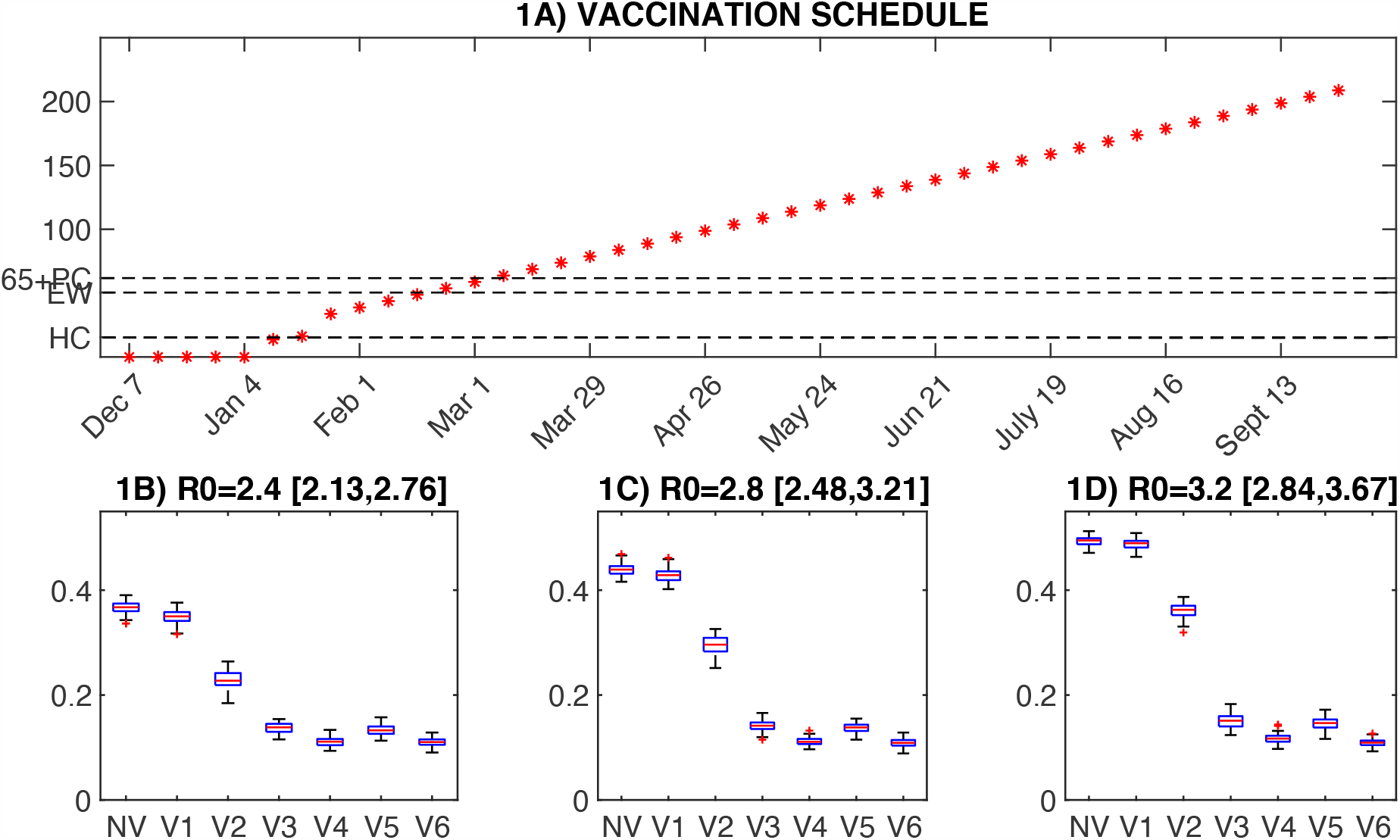
Vaccination schedule and attack rates for different vaccination and NPI scenarios. **1A)** Timeline of vaccination. The prioritization groups: healthcare workers (HC), other essential workers (EW) and 65+ with risk factors (65+PC) were vaccinated subsequently. We set 80%, 60% and 70% target coverages respectively for the 3 prioritization groups, corresponding to 15.5, 35.5 and 11.5 million individuals vaccinated (targets are marked on the y-axes). The vaccine was allocated to the 50 states in proportion to the percentage of residents belonging to each prioritization group. This vaccination schedule was used for simulating scenarios V1 to V6 of panels 1B, 1C and 1D. After the prioritization groups, we allocated vaccine to adults with pre-existing condition (70% target coverage), then children and other adults (60% target coverage). See Table S3 for details on vaccine allocation. **1B**,**1C**,**1D)** Distribution of the total attack rate (red lines are median values, blue boxes are the interquartile, and whiskers extend to the maximum and minimum values that are not outliers) in the US from December 4^th^ to the end of the pandemic for different estimates of R_0_ and different scenario combinations of vaccination and NPIs.

## Results

Figures 1B-1D compare the proportion of US population infected (attack rate) for 7 different combinations of interventions and 3 different estimates of the basic reproductive number (R_0_) in the absence of NPIs. The baseline (NV) scenario with no vaccination and a complete relaxation of NPIs produces a 37-50% attack rate from December 4 through the end of September, depending on the basic reproductive number. With vaccination and the same complete relaxation of NPIs (Scenario V1), the attack rate would be reduced only 1-2%. In contrast, the attack rate is sizably reduced to 11%-15% when NPI controls are maintained until a substantial portion of the population has been vaccinated (Scenarios V3-V6).

## Discussion

Our current estimate of the effective reproductive number (R_t_= [0.8-1.2]) reflects not only the diminished susceptibility of the population due to accumulated natural immunity, but also the reduction in contact rates due to NPIs. However, NPIs are temporary mitigation measures, and their relaxation prior to comprehensive population-scale vaccination will re-inflate R_t_ and, in turn, the threshold for herd immunity. Overall, vaccination uptake over the next 9 months could prevent infection for up to 26% to 39% of the US population; however, this potential benefit depends strictly on maintaining NPIs during vaccine deployment: relaxing NPIs before attaining adequate vaccine coverage could result in tremendous loss of potentially averted cases, hospitalizations and mortality. In the limit scenario V1 in which all NPIs are immediately relaxed before the vaccination campaign, the averted infections are nominal. The findings based on this rapid analysis underscore the importance of maintaining NPIs throughout the upcoming SARS-CoV-2 vaccination campaign to maximize the public health benefit; more in-depth modeling will be performed as more information becomes available on vaccine availability.

Public health messaging is critically needed to encourage continued compliance with NPI control measures in the coming months.

## Data Availability

All model output are available upon request.

## SUPPLEMENTARY MATERIAL

### 1 Model Description

The model is a single location Susceptible-Exposed-Infected-Recovered-Vaccinated (SEIRV) compartmental structure run in isolation for each state in the US. We accounted for 5 age groups and 4 population types: healthcare workers (HC), essential workers (EW), individuals with pre-existing conditions (PC) and others (O) (see Table S1). This compartmental model is a modification of the structure that our group has used for inference and forecast for multiple infectious diseases, including influenza [Shaman, 2012].

**Table S1:** Population stratification.

|  | POPULATION GROUP |
| --- | --- |
| 1 | 0-4 |
| 2 | 5-17 |
| 3 | 18-49 O |
| 4 | 18-49 HC |
| 5 | 18-49 EW |
| 6 | 18-49 PC |
| 7 | 50-64 O |
| 8 | 50-64 HC |
| 9 | 50-64 EW |
| 10 | 50-64 PC |
| 11 | 65+ O |
| 12 | 65+ PC |

The complete stratified model for each state s=1:50 reads:

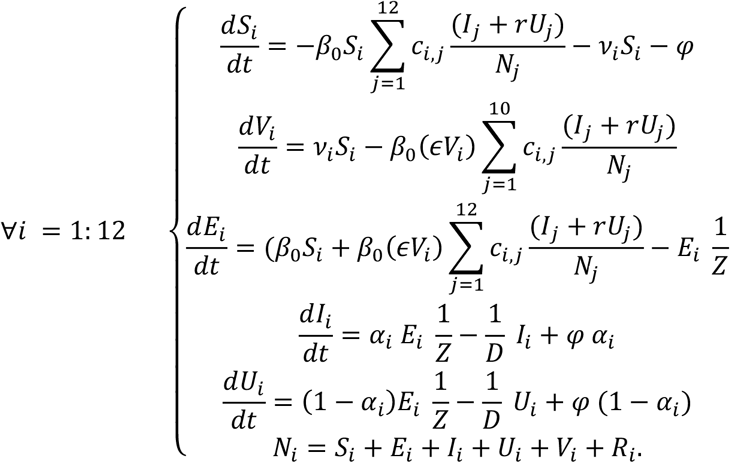

For each group *S*_*i*_, *E*_*i*_, *I*_*i*_, *U*_*i*_, *V*_*i*_ and *R*_*i*_ represent the susceptible, exposed, infected reported and unreported, vaccinated and recovered populations, *D* the duration of infectious period, Z the latency period, *N* the population size, *α*_*i*_ the ascertainment rate of infection, *φ* travel-related importation of SARS-COV-2 into the model domain, *v*_*i*_ the vaccination rate, and *ε* vaccine (in)effectiveness. We allow the transmission rate to vary through specific age-dependent contact rates *c*_*i,j*_ between individuals in age group N_i_ and N_j,_ such that the transmission term reads 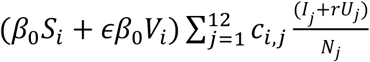. Parameter *r* reflects the decreased probability of transmission for unreported infectious individuals. In the present model, we did not consider waning immunity and the possibility of reinfection.

The model was parametrized based on inference methods described in Section 2, and 100 ensemble projections were simulated in each state for 400 days starting from initial conditions estimated on December 4^th^.

### 2 Parametrization

Our strategy for defining the distributions of parameters and variables in system (1) is based on the following steps:

1. We estimated the *population-level* distribution (interquartile range) of the epidemiological parameters in each State with a non-stratified metapopulation model/ EAKF filter framework. Details on the inference procedure can be found in [Pei, 2020].
2. We combine the *population-level* estimates with additional data and published estimates to stratify the parameters by age and population type (see Table S2 for details on specific parameters and variables).
3. We initialized system (1) with the stratified distribution of parameters and initial conditions. For each state, we ran 100 simulations, each with initial conditions and parameters randomly drawn from the estimated distributions.

**Table S2:** Age- and population-specific parameter specifications. Although all parameters and variables are state-specific, we omitted the subscript _S=1,,50_ for the sake of clarity. *Estimated* parameters were inferred with the model-inference framework described in [Pei, 2020] using incidence data at the County level up to December 4^th^.

| PARAMETER | ESTIMATED | ADDITIONAL DATA | MODEL |
| --- | --- | --- | --- |
| Population structure $N_i$ by state | | Census population structure for 50 states divided into the 5 age groups. [US CENSUS]<br>[CDC, People at Increased risk] | - 40% adults are EW, among them 25% are HC<br>- 30% of the remaining adults have health conditions.<br>- 39% of the 65+ age group have health conditions. |
| State Age-specific transmission rates $\beta_{i,j}$ | State specific $R_0$ (non-stratified population) | *Polymod Study* [Prem, 2017] contact rates at home, school, work, others.<br>CDC modeling assumption: EW are able to reduce work contact 35% as much as the others [ACIP, CDC] | We define the transmission rate $\beta_{i,j}$ so that the reproductive number matches the estimated $R_0$ from Dec 4<sup>th</sup><br>**Contact Matrix $M = \{c_{i,j}\}$:**<br>Contact reductions to account for NPIs: 60% in school, 60% work, 30% others.<br>EW work contact reductions: 20%<br>Contacts of every group with adult group 18-45 and 45-64 are distributed within the 4 adult subgroups (HC, EW, PC, O) according to prevalence.<br>**Transmission rate:**<br>$\beta_{i,j} = \beta_0 c_{i,j}$<br>where $\beta_0$ is determined for each State by equating the $R_0$ computed from the next generation matrix of system (1) to the $R_0$ inferred with model in [2]. |
| Ascertainment rates by age $i$ and state $S$: $\alpha_i$ | $\alpha$ non-stratified | Seroprevalence by age and State $p_i$ (latest data from end of September) *Nationwide Commercial Laboratory Seroprevalence Survey* [CDC]<br>Estimate $\alpha_{i,NY}$ of ascertainment rate by age group in NYC (estimate from November 15<sup>th</sup>) [Yang, 2020] | We set the age-specific ascertainment rate proportional to Yang's estimate:<br>$\alpha_i = k \alpha_{i,NY}$<br>where $k$ verifies: $\alpha = \frac{\sum_i \alpha_i p_i N_i}{\sum_i p_i N_i}$ and $N_i$ and $p_i$ are the population of age group $i$ in state $S$ and the seroprevalence in state $S$.<br>The same ascertainment rate is used for different categories of the same age group, with the exception of the subgroup with pre-existing conditions, which has rate $2\alpha_i$. |
| Initial Susceptibility by age $i$ and state $S$: $S_{0,i}$ | $S_0$ non-stratified | Seroprevalence by age and State $p_i$ (latest data from end of September) | We set $S_{0,i}$ so that the sum over the groups is equal to the estimated $S_0$ on Dec 4<sup>th</sup> and the relative ratio of infection |
| | | *Nationwide Commercial Laboratory Seroprevalence Survey [CDC]* | prevalence across age group matches the serological observations:<br>$S_{0_i} = N_i - u p_i N_i$<br>where $u$ verifies<br>$N - S_0 = u \sum p_i N_i$<br>and $N_i$ and $p_i$ are the population of age group $i$ in state $S$ and the seroprevalence in state $S$.<br>(The same seroprevalence was adopted for different groups of same age). |
| Initial number of infected individuals by age and state:<br>$E_{0_i}, I_{0_i}, U_{0_i}$ | $E_0, I_0, U_0$ non-stratified | Seroprevalence by age and State $p_i$ (latest data from September 30<sup>th</sup>) | We set initial condition for $E_{0_i}, I_{0_i}, U_{0_i}$ to match the seroprevalence ratio across age groups, the sum over the groups matches the posterior estimates for $E_0, I_0, U_0$ on Dec. 4<sup>th</sup>:<br>$E_{0_i} = h_i E_0$<br>where $h_i = \frac{p_i N_i}{\sum_i p_i N_i}$<br>and similarly, for $I_{0_i}, U_{0_i}$. |
| Duration of incubation $Z$ | $Z$ estimated | | Assumed constant for all groups |
| Duration of infectious period $D$ | $D$ estimated | | Assumed constant for all groups |
| Relative infectiousness $r$ of unreported cases | $r$ estimated | | Assumed constant for all groups |

### 3 Vaccine deployment

Two vaccines (I1 and I2) were included in the simulated vaccination campaign. We assumed 20 million doses of I1 available the week of December 14 and 20 million doses of I2 available the week of December 21. For each vaccine, 5 million doses were additionally available each subsequent week. Both vaccines required 2 doses and complete efficacy (90%) was reached 1 week after the second dose. We did not account for partial immunity between doses. We assumed that each individual received the second dose exactly 3 (I1) and 4 (I2) weeks after the first dose. Weekly doses were distributed uniformly over 7 days. Vaccine distribution timeline is detailed in Table S3.

**Table S3:** Vaccine distribution timeline (in millions) for vaccines I1 and I2. *Vacc. Total* denotes the number of individuals that have received the second dose by > 1 week.

| WEEK | 1<sup>ST</sup><br>DOSE<br>I1 | 2<sup>ND</sup><br>DOSE<br>I1 | VACC.<br>V1 | TOT<br>DOSES<br>I1 | 1<sup>ST</sup><br>DOSE<br>I2 | 2<sup>ND</sup><br>DOSE<br>I2 | VACC.<br>V2 | TOT<br>DOSES<br>I2 | VACC.<br>TOTAL | VACC/<br>WEEK |
| --- | --- | --- | --- | --- | --- | --- | --- | --- | --- | --- |
| 14-Dec | 13.7<br>5 | 0 | 0 | 13.7<br>5 | 0 | 0 | 0 | 0 | 0 | 0 |
| 21-Dec | 2.5 | 0 | 0 | 16.2<br>5 | 15 | 0 | 0 | 15 | 0 | 0 |
| 28-Dec | 2.5 | 0 | 0 | 18.7<br>5 | 2.5 | 0 | 0 | 17.5 | 0 | 0 |
| 4-Jan | 2.5 | 13.7<br>5 | 0 | 35 | 2.5 | 0 | 0 | 20 | 0 | 0 |
| 11-Jan | 2.5 | 2.5 | 13.75 | 40 | 2.5 | 0 | 0 | 22.5 | 13.75 | 13.75 |
| 18-Jan | 2.5 | 2.5 | 16.25 | 45 | 2.5 | 15 | 0 | 40 | 16.25 | 2.5 |
| 25-Jan | 2.5 | 2.5 | 18.75 | 50 | 2.5 | 2.5 | 15 | 45 | 33.75 | 17.5 |
| 1-Feb | 2.5 | 2.5 | 21.25 | 55 | 2.5 | 2.5 | 17.5 | 50 | 38.75 | 5 |
| 8-Feb | 2.5 | 2.5 | 23.75 | 60 | 2.5 | 2.5 | 20 | 55 | 43.75 | 5 |
| 15-Feb | 2.5 | 2.5 | 26.25 | 65 | 2.5 | 2.5 | 22.5 | 60 | 48.75 | 5 |
| 22-Feb | 2.5 | 2.5 | 28.75 | 70 | 2.5 | 2.5 | 25 | 65 | 53.75 | 5 |
| 1-Mar | 2.5 | 2.5 | 31.25 | 75 | 2.5 | 2.5 | 27.5 | 70 | 58.75 | 5 |
| 8-Mar | 2.5 | 2.5 | 33.75 | 80 | 2.5 | 2.5 | 30 | 75 | 63.75 | 5 |
| 15-Mar | 2.5 | 2.5 | 36.25 | 85 | 2.5 | 2.5 | 32.5 | 80 | 68.75 | 5 |
| 22-Mar | 2.5 | 2.5 | 38.75 | 90 | 2.5 | 2.5 | 35 | 85 | 73.75 | 5 |
| 29-Mar | 2.5 | 2.5 | 41.25 | 95 | 2.5 | 2.5 | 37.5 | 90 | 78.75 | 5 |
| 5-Apr | 2.5 | 2.5 | 43.75 | 100 | 2.5 | 2.5 | 40 | 95 | 83.75 | 5 |
| 12-Apr | 2.5 | 2.5 | 46.25 | 105 | 2.5 | 2.5 | 42.5 | 100 | 88.75 | 5 |
| 19-Apr | 2.5 | 2.5 | 48.75 | 110 | 2.5 | 2.5 | 45 | 105 | 93.75 | 5 |
| 26-Apr | 2.5 | 2.5 | 51.25 | 115 | 2.5 | 2.5 | 47.5 | 110 | 98.75 | 5 |
| 3-May | 2.5 | 2.5 | 53.75 | 120 | 2.5 | 2.5 | 50 | 115 | 103.75 | 5 |
| 10-May | 2.5 | 2.5 | 56.25 | 125 | 2.5 | 2.5 | 52.5 | 120 | 108.75 | 5 |
| 17-May | 2.5 | 2.5 | 58.75 | 130 | 2.5 | 2.5 | 55 | 125 | 113.75 | 5 |
| 24-May | 2.5 | 2.5 | 61.25 | 135 | 2.5 | 2.5 | 57.5 | 130 | 118.75 | 5 |
| 31-May | 2.5 | 2.5 | 63.75 | 140 | 2.5 | 2.5 | 60 | 135 | 123.75 | 5 |
| 7-Jun | 2.5 | 2.5 | 66.25 | 145 | 2.5 | 2.5 | 62.5 | 140 | 128.75 | 5 |
| 14-Jun | 2.5 | 2.5 | 68.75 | 150 | 2.5 | 2.5 | 65 | 145 | 133.75 | 5 |
| 21-Jun | 2.5 | 2.5 | 71.25 | 155 | 2.5 | 2.5 | 67.5 | 150 | 138.75 | 5 |
| 28-Jun | 2.5 | 2.5 | 73.75 | 160 | 2.5 | 2.5 | 70 | 155 | 143.75 | 5 |
| 5-Jul | 2.5 | 2.5 | 76.25 | 165 | 2.5 | 2.5 | 72.5 | 160 | 148.75 | 5 |
| 12-Jul | 2.5 | 2.5 | 78.75 | 170 | 2.5 | 2.5 | 75 | 165 | 153.75 | 5 |
| 19-Jul | 2.5 | 2.5 | 81.25 | 175 | 2.5 | 2.5 | 77.5 | 170 | 158.75 | 5 |
| 26-Jul | 2.5 | 2.5 | 83.75 | 180 | 2.5 | 2.5 | 80 | 175 | 163.75 | 5 |
| 2-Aug | 2.5 | 2.5 | 86.25 | 185 | 2.5 | 2.5 | 82.5 | 180 | 168.75 | 5 |
| 9-Aug | 2.5 | 2.5 | 88.75 | 190 | 2.5 | 2.5 | 85 | 185 | 173.75 | 5 |
| 16-Aug | 2.5 | 2.5 | 91.25 | 195 | 2.5 | 2.5 | 87.5 | 190 | 178.75 | 5 |
| 23-Aug | 2.5 | 2.5 | 93.75 | 200 | 2.5 | 2.5 | 90 | 195 | 183.75 | 5 |
| 30-Aug | 2.5 | 2.5 | 96.25 | 205 | 2.5 | 2.5 | 92.5 | 200 | 188.75 | 5 |
| 6-Sep | 2.5 | 2.5 | 98.75 | 210 | 2.5 | 2.5 | 95 | 205 | 193.75 | 5 |
| 13-Sep | 2.5 | 2.5 | 101.25 | 215 | 2.5 | 2.5 | 97.5 | 210 | 198.75 | 5 |
| 20-Sep | 2.5 | 2.5 | 103.75 | 220 | 2.5 | 2.5 | 100 | 215 | 203.75 | 5 |
| 27-Sep | 2.5 | 2.5 | 106.25 | 225 | 2.5 | 2.5 | 102.5 | 220 | 208.75 | 5 |

Vaccines were distributed according to the following prioritization schedule (doses required and target coverage)

1. Healthcare workers (31 million doses necessary to fully immunize 80%)
2. Essential workers (70 million doses necessary to fully immunize 60%)
3. 65+ with pre-existing conditions: (22.5 million doses necessary to fully immunize 70%)
4. Other 65+ (70%)
5. Adults with pre-existing conditions (70%)
6. Children 1-4 y/o, followed by children 5-17 (60%); note, the vaccines are currently not indicated for children.
7. Other adults (60%)

